# An Evaluation of Age-varying Genetic Effects underlying Body-mass Index and Blood Pressure in the UK Biobank

**DOI:** 10.64898/2025.12.16.25342301

**Authors:** Genevieve M Leyden, Panagiota Pagoni, Grace M. Power, David Carslake, Tom G Richardson, Kate Tilling, Gibran Hemani, George Davey Smith, Eleanor Sanderson

**Affiliations:** MRC Integrative Epidemiology Unit (IEU), Population Health Sciences, Bristol Medical School, University of Bristol, Oakfield House, Bristol, BS8 2BN, United Kingdom; Centre for Neuropsychiatric Genetics and Genomics, Division of Psychological Medicine and Clinical Neurosciences, School of Medicine, Cardiff University, United Kingdom; Institute for Molecular Bioscience, University of Queensland, Brisbane, Australia

**Author notes:** Corresponding author: Genevieve M Leyden, MRC Integrative Epidemiology Unit (IEU), Population Health Sciences, Bristol Medical School, University of Bristol, Oakfield House, Bristol, BS8 2BN, United Kingdom.

## Abstract

Genome-wide association studies (GWAS) are conventionally conducted in cohorts spanning a wide age-range. These studies typically assume that genetic associations are constant across different ages. Some traits, however, may have age-varying genetic associations. This has implications for the interpretation of genetic effects derived in downstream applications, such as Mendelian randomization (MR) analyses. In this study we conducted a series of age-stratified GWAS on individuals aged 40-69 years in the UK Biobank, for body-mass index (BMI) and three blood pressure traits (systolic, diastolic and pulsatile pressure (PP)) in 2-year age strata (N up to 26,330). We used a meta-regression approach to systematically identify single nucleotide polymorphisms (SNPs) with evidence for age interaction effects among trait-associated GWAS signals and additional loci genome-wide. Within an MR framework, we examine the relationship between BMI and blood pressure traits on cardiovascular and cardiometabolic outcomes (type-2 diabetes (T2D), stroke, peripheral artery disease (PAD), heart failure, coronary heart disease and atrial fibrillation). Next, we describe the effect of the SNP^*^Age interaction on those relationships in a modified inverse-variance weighted (*ivw*) analysis. We identified differential enrichment of age-interaction effects, which was trait dependent. For example, 10.3% of BMI discovery SNPs had evidence for an age-interaction in our data compared to 44.7% for PP (at P<0.05). Our downstream MR and modified *ivw* analyses highlight the influence of age on the genetically predicted relationship between PP and adverse cardiovascular outcomes. For example, our results indicated that an increased rate of change in genetically predicted PP across the age period is associated with higher susceptibility to PAD (interaction odds ratio= 2.71; P=1.82×10^-13^; 95%-CI: 2.08-3.53). The data generated in this project provides a valuable resource for further exploration of mechanisms relevant to the genetic architecture of complex traits and all summary data will be made readily accessible to the research community.

**Author Summary:** Genetic variants which reliably predict variation in a trait are a valuable tool within genetic epidemiology studies, offering a means to estimate whether an exposure-outcome relationship is likely to be causal using a method called Mendelian randomization (MR). Typically, MR results are interpreted as the cumulative lifetime effect of the exposure on the outcome. However, there is growing evidence which suggests that the influence of genetic effects on trait variation detected in cross-sectional population studies may be age dependent in some scenarios. In this work we aimed to conduct a thorough investigation on whether and to what extent the influence of genetics on population-level trait variation changes across adulthood. We investigated this question within a methodological framework which used age-stratified summary level data, demonstrating that this approach may have wide applicability to the research community where individual level cohort data are not publicly available. We demonstrate that age interacts with genetic influences across adulthood in a trait dependent manner, where genetics may have a stronger influence on variation in body-mass index measured earlier in life, and on pulsatile pressure later in life. We take advantage of the MR and *ivw* frameworks to further illustrate how the variation in the exposure explained by genetics varies with increasing age. This exploratory work helps provide insight on the extent that distinct genetic effects are detectable across adulthood, helping us to understand how more precise lifecourse effects may be genetically proxied within an MR setting.

## Introduction

Genome-wide association studies (GWAS) are typically conducted in large cohorts representing age-diverse adult participants where it is assumed that the genetic associations are constant across the sample age-range. However, there are many examples of age-dependent genetic associations with complex traits which are not typically captured by standard cross-sectional study designs (1-4). The genetics of body-size is a noteworthy example, as genetic predictors of childhood, early-mid adulthood, and mid-later adulthood body-size vary considerably and have been shown to independently influence later-life disease risk (5-7). GWAS conducted on childhood cohorts including children aged 2-10 years have also identified distinct genetic signals on body-size compared with adulthood (8), while a number of age-dependent genetic effects have been identified on blood pressure traits between early and late-adulthood within age-stratified GWAS initiatives (3). The contribution of population level variance quantitative trait loci (vQTL) to the genetic architecture of anthropometric traits in particular may also be substantially influenced by genotype^*^age interactions (9, 10), where the trajectory of within-individual trait variation has also been shown to be driven by age (11). Age-varying genetic associations may have important implications within the context of Mendelian randomization (MR) analyses, where genetically instrumented exposures are typically interpreted as the lifetime effect of the exposure on an outcome (12-14). Given the prevalence of chronic conditions such as high blood pressure and obesity and their subsequent relationship with disease onset, a better understanding of how the genetic architecture of complex traits varies by age may provide important insight into the biology and temporal effects underlying risk.

The UK Biobank (UKB), comprising health and genotype data on >450,000 participants aged between 40-69 years, offers a valuable platform to investigate age-dependent genetic effects in an adult population (15). Previous studies have aimed to estimate putative age-dependent effects underlying variation in blood pressure on disease risk in an MR setting. In these examples, the UKB cohort was separated into groups aged younger or older than 55 years. Genetic instruments for blood pressure traits were then identified in each group and incorporated as separate exposures using a multivariable MR framework (16, 17). However, estimation of distinct effects of the exposure in each age group on an outcome in a multivariable MR setting is dependent on there being variation in the genetic effect predicting the exposure in each age group (18-20). Some studies have shown that the extent to which genetic variants predict trait variation diminishes with increasing age (1). As such, variation in genetic effects with age will have implications for the gene-environment equivalence principle of MR, which posits that modifications in the exposure which are due to either genetic or environment effects, will have an equivalent effect on the outcome. This may be partly attributable to apparent age-varying trait associations arising by reverse causation due to prodromal disease, or they may arise indirectly through the effect of disease on study participation (21). For example, a missense variant in the *APOE* gene (rs429358) is both one of the strongest genetic risk factors for Alzheimer’s disease (AD) (22) and has been shown to strongly associate with decreases in body-mass index (BMI) and weight with increasing age (23). While AD liability has long been associated with a variety of lifestyle and biological phenotypes (including lower BMI) (24, 25), evidence from MR studies has indicated that this is unlikely to be causal but potentially driven by selection or reverse cause (26, 27). Notably, genetic variants at *APOE* (implicated in AD) and *CHRNA3* loci (implicated in smoking heaviness) have produced consistent effects on survival to late age (28), with smoking being a major contributor to premature cardiovascular related death (29). For these reasons further in-depth investigation of the extent of age-dependent genetic effects is warranted.

In this study, we aimed to comprehensively evaluate evidence of age-dependent genetic effects across the UKB cohort on blood pressure traits (pulsatile pressure (PP), systolic blood pressure (SBP), diastolic blood pressure (DBP)), and BMI within an age-stratified framework. An overview of our analytical framework is provided in **Figure 1**. While all three blood pressure traits are included in the study, we are primarily interested in the age-varying genetic associations underlying PP. By capturing the difference between SBP and DBP, an elevated PP is a useful hallmark of an aging cardiovascular system (52). PP is also commonly used as a proxy indicator of vascular stiffness and is an important predictor of adverse cardiovascular outcomes (53).

**Figure 1.**
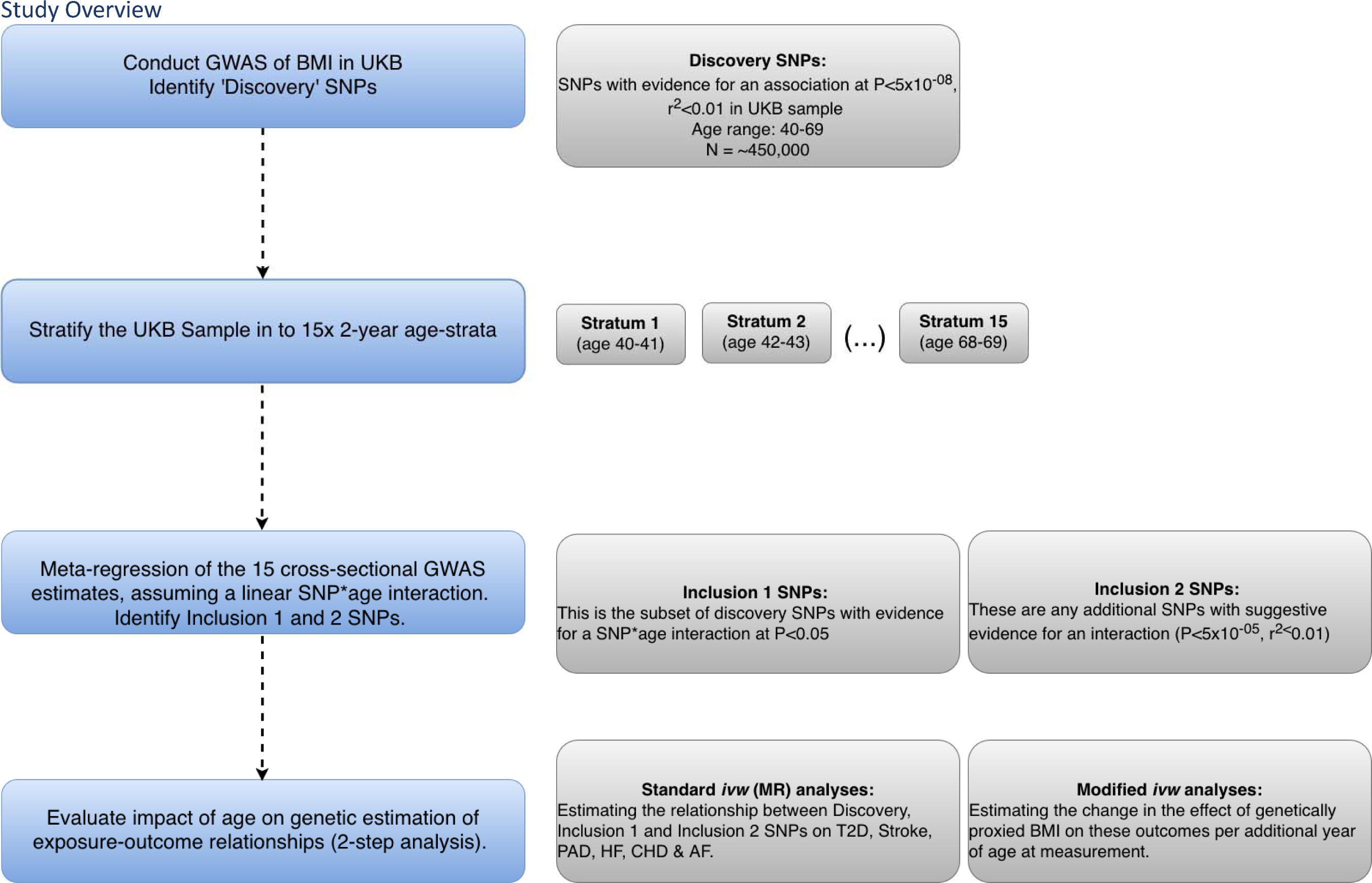
Study overview outlining process for one trait (BMI as exemplar). Discovery SNPs were first identified across the full UKB cohort aged 40-69 years. The UKB cohort was then stratified into 15 two-year age strata, and individual GWAS analyses were conducted within each stratum for all traits of interest (BMI, PP, SBP, DBP). Evidence for age-varying genetic effects was next evaluated by meta-regression analysis. We employed 2 strategies to identify loci with putative age-varying genetic effects. First, our inclusion 1 set of variants are the subset of discovery SNPs with evidence for an age-varying effect in the meta-regression analysis at P<0.05. Secondly, our inclusion 2 SNPs evaluated all loci genome-wide with evidence for an effect at P<5×10^-05^ and r^2^ <0.01. Finally, the SNPs identified by our inclusion criteria were used in ivw analyses using Mendelian randomization to evaluate the relationship between each set of variants and secondary outcomes (standard ivw), and the influence of age on the estimation of the relationship (modified ivw) using genetic methods.

A major motivation for conducting GWAS in an age-stratified setting was to develop an analytical framework based on summary level data which could have wide utility for the inclusion of additional cohorts where individual level data is often not available. In addition, by a generating a dataset comprising age-specific GWAS summary statistics where no assumption on the shape of the relationship with age is imposed on the dataset generated, this resource may have utility in detecting SNPs with non-linear peaks in association by age. To achieve this, separate GWAS were carried out for each trait on the UKB cohort stratified into 15 two-year age strata, and on a ‘discovery’ dataset comprising the full cohort (aged 40-69 years). Additionally, we conducted an individual-level genome-wide SNP^*^Age interaction analysis on the sample comprising the full age-range as a sensitivity analysis. Lastly, to formally investigate evidence for SNP^*^Age interactions, meta-regression was used to model a linear relationship between the age-stratified GWAS results and age. Meta-regression approaches have recently been demonstrated to appropriately model SNP^*^Age interaction effects when age-specific GWAS summary results are available (3, 30-32). The relationship between the SNP^*^Age interactions identified for each trait and cardiovascular and cardiometabolic outcomes were subsequently evaluated within an MR and modified *ivw* framework, providing insight on the changing influence of genetic effects on the rate of change in the measured exposure across the 40-69 year age-range. This study provides an applied example of the use of meta-regression as a platform for the systematic identification of context-dependent genetic effects and an evaluation of their prevalence with respect to complex traits.

## Results

### Identification of “discovery” GWAS signals in individuals aged 40-69 years

GWAS associations were initially generated for each trait (BMI, PP, SBP and DBP) on imputed genetic data for individuals aged 40-69 years in UKB (N∼450,000), which will herein be referred to as the ‘discovery’ GWAS. The estimated variance explained in our discovery GWAS was 6.8%, 3.8%, 4.7% and 4.8% for BMI, PP, SBP, and DBP, respectively (and in line with previous studies (33, 34), though likely inflated due to winner’s curse. Our ‘discovery’ GWAS analyses (on the 40-69 year age range) identified 1,150, 795, 888, and 880 independent SNPs associated with BMI, PP, SBP, and DBP, respectively, based on a genome-wide significance threshold (P<5×10^-8^ ) and an r^2^ <0.01 for independence.

### Age-stratified GWAS results

We next generated a set of GWAS results on the sample of 40-69 year olds in the UKB stratified into 15 two-year age strata for each trait (stratum 1: 40-41 year olds; stratum 2: 42-43 year olds etc.). A feature of the age-stratified resource we have generated is that a linear constraint is not imposed on overall dataset. Therefore, in our preliminary investigation of the age-stratified GWAS results we aimed to visualize and assess any potential trends, or if peaks in trait-associations may occur at middle-ages by plotting the SNPs with the largest differential effect between selected age-groups.

As expected, detection of genome-wide significant trait effects within 2-year age strata was low due to reduced power following stratification (tables outlining the lead variants identified in each individual age-stratified GWAS are provided in **Table S1-4**). As such, our initial investigation of the results focus on the trait associated SNPs identified in our “discovery” GWAS on the sample aged 40-69 years (we refer to as “discovery SNPs”) and how their effect estimates compare between age-strata specific GWAS results. Visual comparisons of the effect estimates derived for discovery SNPs between distant age-strata are presented in **Supplementary Material** (**Figures S1-S12**).

All GWAS results generated as part of this study are currently under restricted access on Zenodo. Access to this dataset will be made publicly available upon acceptance of this work for publication.

### Identification of SNP^*^Age interaction effects by meta-regression

Next, we aimed to explore how the age-stratified GWAS datasets may be used to examine the interactions between age and the SNP effects on the trait. To formally evaluate the extent of linear age-varying genetic association across this age-range, linear SNP^*^Age interaction effects were estimated by meta-regression on a per SNP basis. In this setting, a positive interaction indicates an overall amplification, and a negative interaction indicates overall attenuation, of the genetic effect on the trait with increasing age. We applied inclusion criteria to the results aiming to answer two questions: (1) to what extent do SNP effects identified in a standard cross-sectional GWAS vary by age, and (2) can we detect additional putative age-sensitive SNP-trait associations which did not reach genome-wide significance in the standard cross-sectional GWAS study.

First, we assessed which of the discovery SNPs showed evidence (under a nominal P<0.05 threshold) of a SNP^*^Age interaction in our meta-regression analysis, which is defined as our “Inclusion 1” set (see the Methods section for further detail). Approximately 10% of discovery SNPs were shown to have evidence of an age-varying effect (at P<0.05) for BMI (119/1150 SNPs; 10.3%) while 44.7% of SNPs had evidence for an age-varying effect for PP (355/795 SNPs). SBP and DBP each also showed age-varying effects for approximately 15% of the discovery SNPs, (157/888 SNPs; 17.7% and 122/880 SNPs; 13.9% respectively). Applying a more stringent Bonferroni corrected P-value threshold (0.05/N-discovery SNPs) highlighted the top 3 SNPs for BMI, 5 for PP, 4 for SBP and 5 for DBP. A detailed overview of all results is provided in **Tables S5-8**.

Plots illustrating the SNPs with the strongest (based on lowest P-value) interaction with age are provided in **Figure 2** for BMI and PP. The effect direction in this context is determined by the choice of effect allele and is therefore less informative. Interpretation of interaction effects should instead focus on the nature of its behaviour, i.e. whether the association is going towards the null or away from the null with increasing age. For example, the present analysis provided strong evidence for the rs6751993 (*TMEM*) association with BMI to attenuate towards the null with increasing age (age 40-41 Beta=-0.348, SE=0.077, P=5.7×10^-06^ ; Interaction Beta= 0.009, SE=0.002, P=6.97×10^-07^ ). Similarly, rs11642015 (*FTO*) presented strong evidence for an effect which attenuated towards the null with increasing age (age 40-41 Beta=-0.396, SE=0.058, P=8.9×10^-12^ ; Interaction Beta= 0.007, SE=0.001, P=9.28×10^-07^ ). These examples for BMI highlighted associations that tend towards the null as age increases. While for PP, the variants with the strongest evidence for an interaction with age had effects which got larger with age. For example, rs62481856 (*LINC02577;CCDC71L*) which had the strongest evidence for an increase in effect magnitude on PP with increasing age (age 40-41 Beta=-0.276, SE= 0.128, P=3.2×10^-02^ ; Interaction Beta=-0.041, SE=0.004, p=4.26×10^-23^ ). Strong evidence for an interaction was similarly observed for rs1848050 (*FBN1*) on PP (age 40-41 Beta=0.089, SE=0.172, P=6.1×10-^01^; Interaction Beta=0.05, SE=0.006, P=1×10^-16^ ).

**Fig. 2.** Bubble plots highlighting discovery SNPs which had the strongest evidence of an age-varying effect based on lowest p-value in the meta-regression analysis: (i) and (ii) BMI, (iii) and (iv) PP. ‘Studies’ refers to the SNP effects derived in each age-stratified GWAS study respectively. The size of the bubbles represents each age group’s weight in estimating the SNP^*^Age relationship, where larger bubbles correspond to age groups with more precise effect size estimates. Each age group is plotted at its mean value on the x-axis and the trait effects are plotted on the y-axis.

Secondly, we assessed the evidence of age-varying genetic effects by meta-regression across all SNPs genome-wide (nSNP = 12,321,875, P= 5×10^-8^ and r^2^ <0.01). This was to identify additional SNPs not otherwise captured by the trait associated GWAS SNPs in our discovery set. To facilitate the identification of further putative age-varying genetic signals we evaluated SNPs with evidence for an age varying effect in the meta-regression analysis at P< 5×10^-5^ and r^2^ <0.01, which we refer to herein as our “Inclusion 2” set (see Methods section for more detail). This identified an additional 193 independent SNPs with age-varying effects for BMI, 366 for PP, 244 for SBP and 286 for DBP (meta-regression interaction thresholds: P<5×10^-05^, r^2^ <0.01). The *APOE* variant rs429358 (T allele) exhibited the strongest age-varying signal with BMI in the present analysis, with an effect attenuating towards the null relative to the GWAS Beta in the youngest strata (age 40-41 GWAS Beta= -0.046, SE=0.08, P=0.56); Interaction Beta= 0.009; SE: 0.0018; P=7.44×10^-08^ ). rs12705390 (*LINC02577;CCDC71L*) had the strongest age-interaction signal in the present analysis for PP, with an interaction Beta which aligned directionally with the GWAS Beta observed in the youngest age strata (age 40-41 GWAS Beta= -0.28, SE=0.13, P=0.028; Interaction Beta=-0.04, SE=0.004, P=3.85×10^-23^ ). We additionally highlight SNPs identified after increasing the interaction threshold to the conventional genome-wide significance level, which yielded no variants for BMI, 29 for PP, 1 for SBP and 2 for DBP (meta-regression interaction thresholds: P< 5×10^-8^ and r^2^<0.01). A detailed overview of these results is provided in **Tables S9-12**.

Next, we aimed to gain insight on the characteristics of the SNPs identified through both Inclusion 1 and 2. Scatter plots illustrating the relationship between the SNP-trait associations (provided by the discovery GWAS estimate on the sample aged 40-69 years) and the SNP^*^Age-interaction (identified by the meta-regression analysis) for these SNPs are provided (BMI and PP **Figure 3**; SBP and DBP **Figure S13**). These plots highlight distinct clustering of SNPs identified either by the discovery GWAS only, Inclusion 1 and Inclusion 2 SNPs, for all traits. Opposite trends can be observed for the effect of age on the SNP-trait associations for BMI and PP (**Figure 3**). For BMI, we observe a general tendency for SNP-trait associations to attenuate in effect with increasing age among SNPs identified by our Inclusion 1 and 2 criteria (i.e. SNPs with an age-varying effect), while the SNP-trait associations for the remaining discovery SNPs are stable. In contrast, the SNP-trait associations among Inclusion 1 and 2 SNPs identified for PP tend to have a greater effect with increasing age. A similar relationship is observed for SBP as PP, though the positive trajectory observed among Inclusion 1 SNPs is less pronounced (**Figure S13**). In general, the effect magnitude of Inclusion 2 SNPs appears large. This may be because the trait effect of the majority of Inclusion 2 SNPs provided evidence of a change in direction between age-strata (proportion of Inclusion 2 SNPs where Beta sign changes; BMI = 97%, PP = 97%, SBP = 99%, DBP = 99%), which may have contributed to an overall smaller average effect in the discovery GWAS conducted across the full age range.

**Figure 3.**
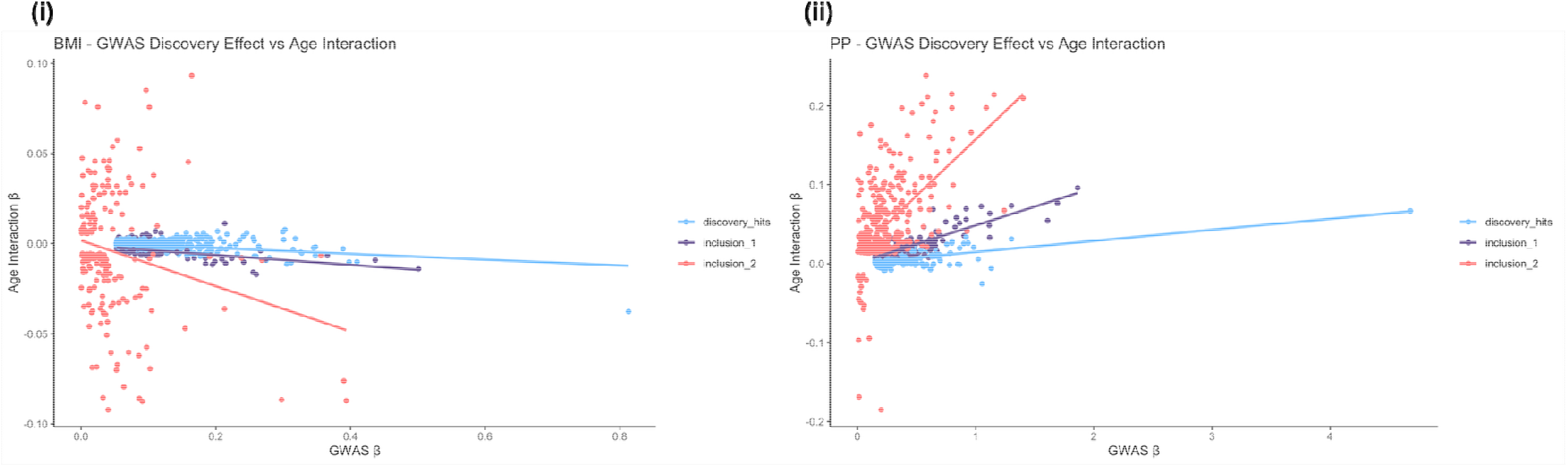
Scatter plot depicting the relationship between SNP-trait effect estimates derived by GWAS and SNP^*^Age interaction effects derived by meta-regression analysis for (i) BMI and (ii) PP. The x-axis depicts the GWAS derived estimates, and the y-axis depicts the meta-regression derived interaction effects.

### Identification of SNP^*^Age interactions in an individual level analysis (sensitivity)

As a comparative study, we conducted genome-wide SNP^*^Age interaction analyses for all traits in a non-stratified individual-level setting (see Methods for detail). Here we observed similar trait dependent trends in the detection of age-varying SNP effects across BMI, PP, SBP and DBP as in the interactions identified by meta-regression. This analysis identified 945 SNPs with evidence for a main trait effect on BMI after clumping (GWAS P<5×10^-08^, r^2^ <0.01), of which 106 had nominal evidence for an interaction with age (SNP^*^Age P<0.05), analogous with our Inclusion 1 criterion in the primary analysis. Additionally, 155 SNPs had evidence for a SNP^*^Age interaction using significance thresholds analogous with our Inclusion 2 criterion (P<5×10^-05^, r^2^ <0.01), of which 4 reached genome-wide significance (P<5×10^-08^ ). The *APOE* variant rs429358 had the strongest evidence for a SNP^*^Age interaction in our individual level analysis (Main effect Beta= 0.056, SE=0.006, P=2.93×10_-20_ ; Interaction Beta=0.0355, SE=0.006, P=3.55×10^-09^ ), consistent with the interaction observed in our meta-regression analysis (Age 40-41 GWAS Beta= -0.046, SE= 0.079, P=0.56; Interaction Beta=0.009, SE=0.002, P=7.44×10^-08^ ).

698 SNPs had evidence for a main effect on PP, with 295 with evidence for an interaction (at SNP^*^Age P<0.05). 267 SNPs had a SNP^*^Age effect at SNP^*^Age P<5×10^-05^, r^2^ <0.01), of which 18 reached genome-wide significance. The strongest SNP^*^Age interaction effect was detected at rs17477177 which maps to the *LINC02577;CCDC71L* locus (Interaction Beta= -0.212, SE=0.02, P= 9.67×10^-22^ ), aligning with the strongest SNP^*^Age interaction detected in our meta-regression which mapped to the same locus. The results of our individual level SNP^*^Age interaction analysis for the remaining blood pressure traits identified 752 SNPs with a main effect for SBP, of which 111 had evidence for an interaction with age at P<0.05, and an additional 183 SNPs were identified across the genome with a SNP^*^Age interaction effect at P<5×10^-05^, r^2^ <0.01 of which none reached genome-wide significance; and 750 main effects for DBP of which 111 had evidence for an interaction with age at P<0.05, and an additional 218 SNPs were identified across the genome with SNP^*^Age interaction effects at P<5×10^-05^, r^2^ <0.01 of which 7 reached genome-wide significance. Further detail on the proportion of overlap between all analyses is provided in **Supplementary Note 2**, and the full details of all variants identified in these analyses are provided in **Tables S13-16**.

### The effect of SNP^*^Age interactions on genetically predicted exposure-outcome relationships

To investigate the impact of age at exposure measurement when using genetically predicted exposures such as in MR, we incorporated our results into a two-step analytical framework. We defined three sets of “instrumental variables” per trait based on the set of discovery, Inclusion 1 and Inclusion 2 SNPs. These sets of variables were incorporated first into a conventional two-sample summary data MR analysis which used the *ivw* method (35) to provide a reference estimate for the effect of the genetically predicted trait (e.g. BMI) on six cardiovascular and cardiometabolic disease outcomes: type-2 diabetes (T2D), stroke, peripheral artery disease (PAD), heart failure (HF), coronary heart disease (CHD) and atrial fibrillation (AF) as predicted by each set of genetic variants. In the second step, we conduct parallel analyses which integrated the SNPs identified by each of our Inclusion criteria into a modified *ivw* analysis. This was achieved using the *ivw* model and substituting the SNP-trait associations (used as instrumental variables in the standard MR) with their corresponding SNP^*^Age interaction effects (see Methods for detail). The resulting modified *ivw* estimates depict the change in the effect of the genetically proxied trait (e.g. BMI) on the outcomes per additional year of age at measurement.

The total genetic effect of BMI as estimated by the discovery set highlighted a strong positive relationship between genetically predicted variation in BMI and the risk of developing each outcome. When estimating effects on traits such as PAD, HF, AF and CHD, the SNPs captured by the Inclusion 1 and Inclusion 2 sets tended to provide less precise estimates of the relationship between BMI and the outcomes evaluated relative to the ‘discovery’ set, while the estimates for stroke attenuated to include the null. BMI strongly predicted risk of developing T2D using the discovery SNPs (odds ratio (OR) = 1.22; 95% confidence interval (CI) = 1.21-1.34; P=1.53×10^-109^ ) and the Inclusion 1 (OR = 1.27; CI = 1.27-1.35; P=5.5×10^-82^ ) and Inclusion 2 sets (OR = 1.27; CI = 1.12-1.34; P=3.59×10^-18^ ) **(Figure 4 (i))**.

**Figure 4.**
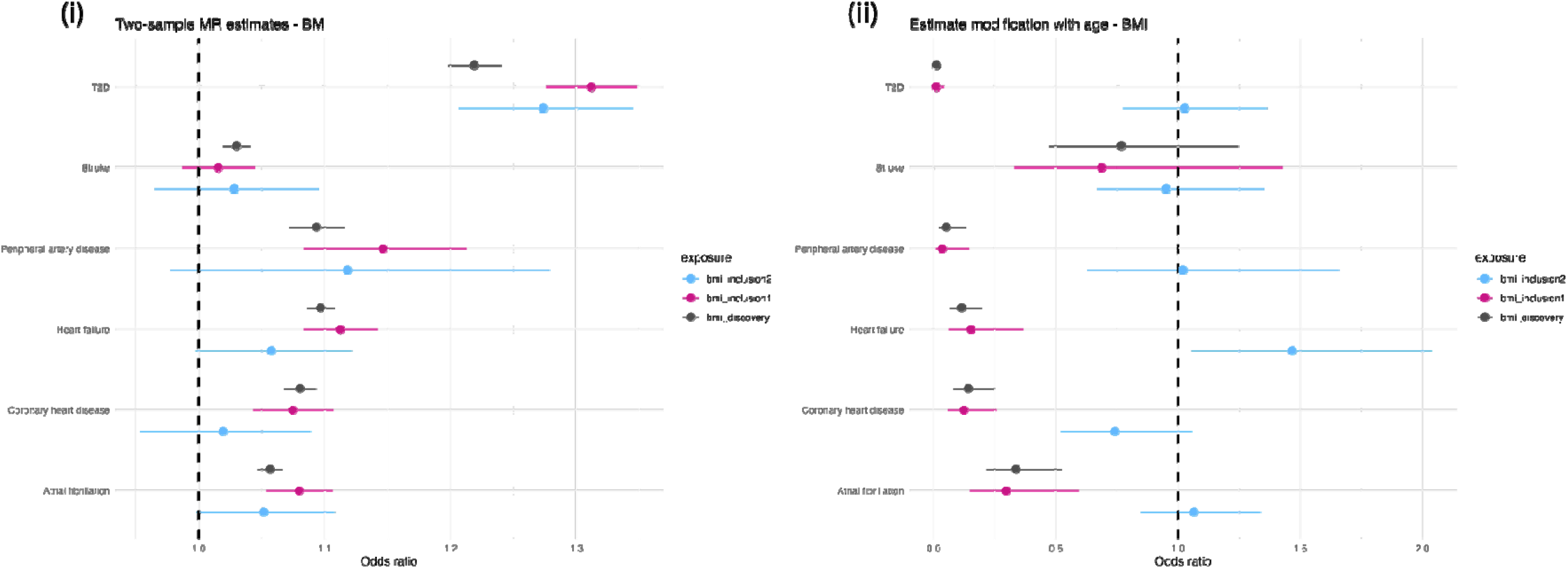
Standard MR estimates of (i) the effect of genetically proxied BMI measured at 40-69 years old on six cardiovascular outcomes and (ii) modified ivw estimates depicting the change in the effect of genetically proxied BMI on those outcomes per additional year of age at measurement. Analyses were conducted separately using SNPs identified by each of our inclusion criteria (discovery, inclusion1 and inclusion2 SNPs) to instrument our exposure variables. Plots depict the log odds estimates (i) per kg m^-2^ increase in BMI and (ii) per kg m^-2^ per year of age. Error bars are 95% confidence intervals.

The standard MR estimates observed for PP on stroke, HF and AF, using SNPs identified through the discovery, Inclusion 1 and Inclusion 2 criteria, were largely similar. In contrast, the Inclusion 2 set for PP predicted a larger OR on PAD (OR = 1.08; CI = 1.06-1.10; P=8.6×10^-13^ ) than the discovery (OR = 1.04; CI = 1.04-1.05; P=5.04×10^-24^ ) or Inclusion 1 set (OR= 1.04; CI = 1.03-1.06; P=1.05×10^-14^ ). This result highlights the potential importance of age of exposure when estimating the genetically predicted effect of PP on PAD **(Figure 5 (i))**.

**Figure 5.**
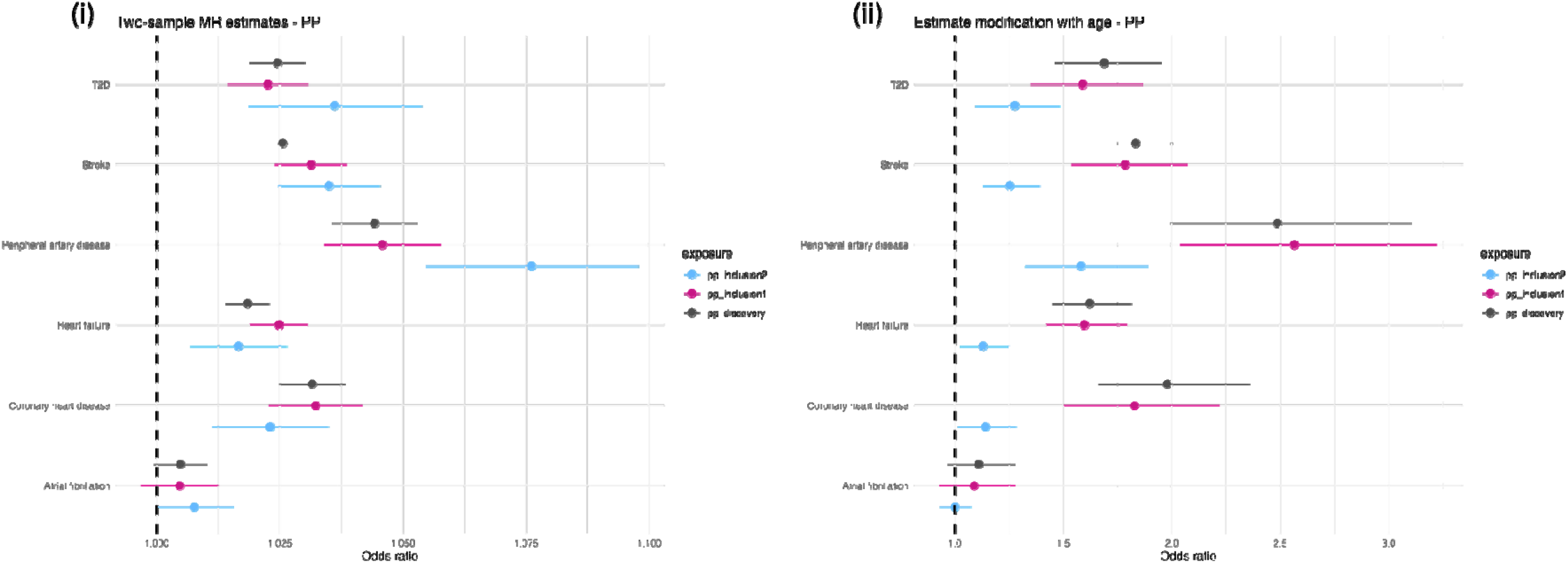
Standard MR estimates of (i) the effect of genetically proxied PP measured at 40-69 years old on six cardiovascular outcomes and (ii) modified ivw estimates depicting the change in the effect of genetically proxied PP on those outcomes per additional year of age at measurement. Analyses were conducted separately using SNPs identified by each of our inclusion criteria (discovery, inclusion1 and inclusion2 SNPs) to instrument our exposure variables. Plots depict the log odds estimates (i) per kg m^-2^ increase in BMI and (ii) per kg m per year of age. Error bars are 95% confidence intervals

Detailed results of standard MR sensitivity analyses using *MR Egger* and *Weighted median* MR methods are provided (**Table S17-S18**). In general, the estimates obtained by standard MR in our analyses of the discovery SNPs agreed in direction of effect across all methods used. We have incorporated the Inclusion 1 and 2 SNPs as instrumental variables in the MR framework to compare how SNPs with evidence for an age-varying trait association perform in the model. As such, these results should not be interpreted as causal effects of the exposure as per conventional MR due to their implications with respect to the gene-environment equivalence assumption of MR. In particular, we note that analyses instrumented using the Inclusion 2 subset of SNPs which are not genome-wide significant for the trait have weaker power to explain trait variation compared to the overall exposure effects instrumented using the discovery SNPs. This is illustrated by examples where an *ivw* result did not replicate across both the weighted median and MR Egger analyses, for example, the *ivw* analysis of BMI-Inclusion 2 SNPs on T2D was replicated by MR Egger (OR=1.31; CI=1.23-1.39; P=3.46×10-14) but not using the weighted median approach (OR= 1.00; CI= 0.94-1.05; P=0.97). The results of additional *ivw* MR analyses comparing the effect estimates for exposures in the youngest and oldest strata are described **Supplementary Note 3** and provided in **Table S19, Figure S14**).

The results of our modified *ivw* analyses illustrate trait dependent differences in the effect of age at measurement on the prediction of exposure-outcome relationships when using genetically derived instrumental variables. For instance, a negative trajectory of association is observed when we consider the effect of increasing age on genetically predicted BMI using the discovery and Inclusion 1 SNP sets on AF, CHD, HF, and PAD, which was most pronounced for T2D (**Figure 4 (ii**)). The effects of the Inclusion 2 SNPs generally centered around the null. Overall, these results suggest that the rate of change in BMI across the 40-69 year age-period which is due to genetic effects appears to associate with more favorable health outcomes. As the effect size of genetic associations for BMI attenuate with increasing age (**Figure 3 (i)**), conceptually this result suggests environmental factors drive a more pronounced rate of change in measured BMI across the same age-period.

The results of the modified *ivw* analysis investigating the impact of age on the genetically predicted relationships between PP and cardiovascular outcomes highlights the opposite trend to that observed for BMI. For example, our analysis identified a strong directionally positive trajectory of association between the rate of change in PP by genetic variation as age increases on CHD, stroke and PAD for our discovery, Inclusion 1 and Inclusion 2 variant sets (**Figure 5 (ii**)). In summary, this indicates that with increasing age in the sample, genetically predicted PP is an increasingly relevant predictor for these outcomes. The results of all modified *ivw* analyses are provided in **Tables S20**. Additional plots depicting the results of analyses investigating SBP and DBP in the modified MR framework are provided in **Figure S15-S16**.

### Examination of bias due to selection in the age-stratified framework

Next, we examined whether age-varying genetic effects identified using an age-stratified framework may be sensitive to sources of selection bias. First, we conducted a simulation study to estimate the potential for selection bias to induce an age-varying genetic effect when the true effect of the SNP on the trait is constant over different ages. The model we consider is;

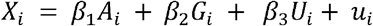

Where *X* is a trait of interest, *A* is an individual’s age, *G* is a genetic variant (e.g. SNP) that influences the trait of interest, U is an unobserved confounder and *U* is a random error term (*u*_*i*_ ∼*N*(0,1)). Selection, i.e. whether or not any of the variables are observed for a particular individual, depends on potentially interacting effects (36) of their age and the level of their trait in the form;

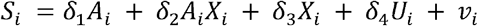

Where *v*_*i*_ is a random error term (*v*_*i*_ ∼*N*(0,1)). *S*_*i*_ is a marker for how likely a variable is to be selected with the top 50% of observations of *S*_*i*_ selected into the dataset for analysis and the other observations removed. The simulated dataset was then stratified into 4 age groups and the association between the genetic variant and the trait was estimated in each group separately. Full details of the simulation are given in the methods section. We varied the values of *δ*_1_, *δ*_2_ and *δ*_3_ to assess the impact of different selection mechanisms on the estimated association in each age group. These results show that when there is no interaction in the effects of age and the trait on selection (i.e. *δ*_2_ = 0), the same association between the genetic variant and the trait is estimated in each age group. However, when there is an interaction, a different effect is estimated in each age group, potentially inducing an apparent age-varying genetic effect (**Figure 6 (i)**). Notably, this age variation in the estimated effect does not occur if there is no effect of the genetic variant on the trait (**Figure 6 (ii)**).

**Figure 6.**
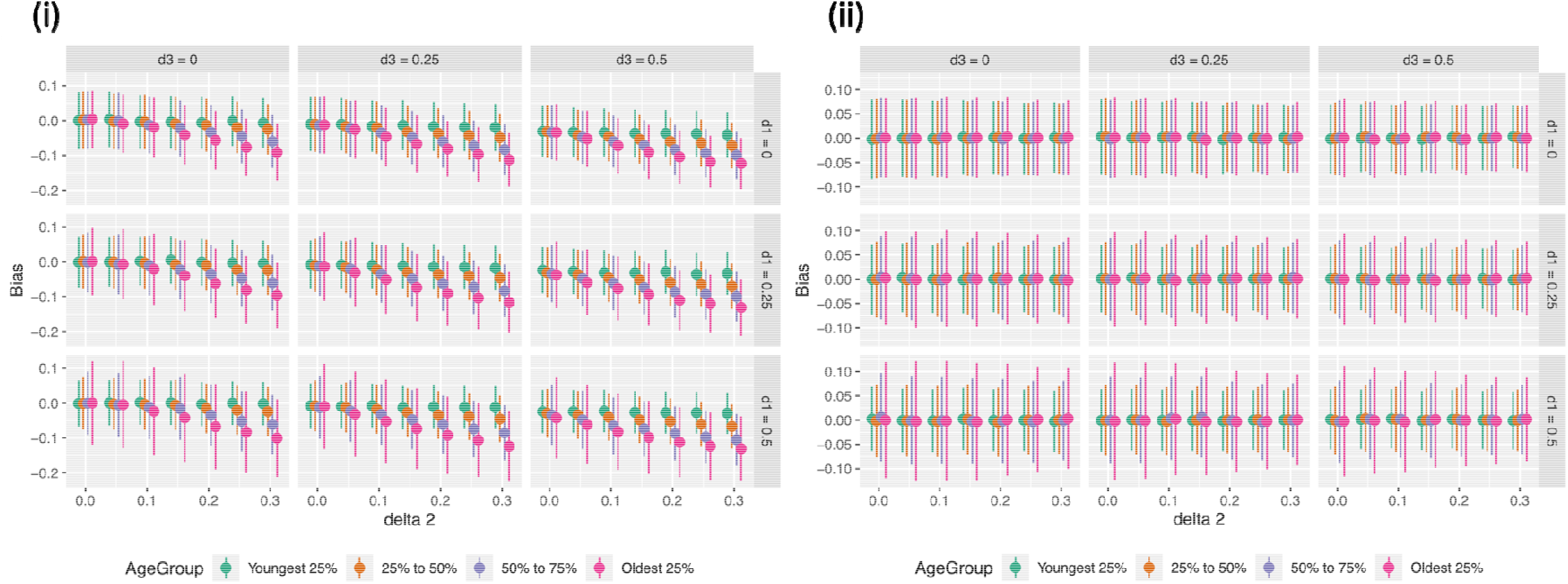
Bias in age-specific estimated genetic effects (i) in the presence of a genetic effect on X and (ii) with no genetic effect on X.

We next explored potential sources of selection bias using an empirical example from the UKB. To do this we evaluated the relationship between BMI and the probability of attending a repeat clinic, in UKB participants stratified into 5 age groups (Figure 7). Dummy variables were created for each age group to examine BMI’s effect on the probability of returning to the clinic without assuming a linear relationship across age groups. Individuals in the youngest age category appeared least sensitive to an effect of BMI on their probability of attending. While older participants appeared more sensitive to such an effect, the observed effects are largely consistent among the older age groups. We conducted further analyses using logistic regression to estimate the relationship between the trait^*^age interaction on the probability of returning to the clinic on each of our traits of interest. Within the youngest age bin, there was some evidence that with increasing age there is a small trait effect on their probability of attending a repeat clinic (Age 39-48 bin Beta=-0.005, SE=0.002, P=0.005). While no evidence for an interaction in the other age bins, the overall estimate indicates that with increasing age, higher BMI may have a small effect on the probability of attending a repeat clinic assessment (Overall Beta: -0.0007, SE=0.0002, P=0.001). Similarly, we found no evidence for an interaction between PP^*^Age within any individual age bin, however, the overall estimate indicated that with age there is strong evidence for a small PP^*^Age interaction effect on the probability of attending a repeat clinic assessment (Overall Beta=-0.0005, SE=0.00007, P=2.84×10^-09^ ). A weaker overall interaction was observed for SBP (Overall Beta=-0.0002, SE=5.43×10^-05^, P=0.003), and DBP (Overall Beta=0.0002, SE=9.4×10^-05^, P=0.048). These results yielded evidence of a small interaction between the trait and age on the probability of turning up (**Table S17**).

**Figure 7.**
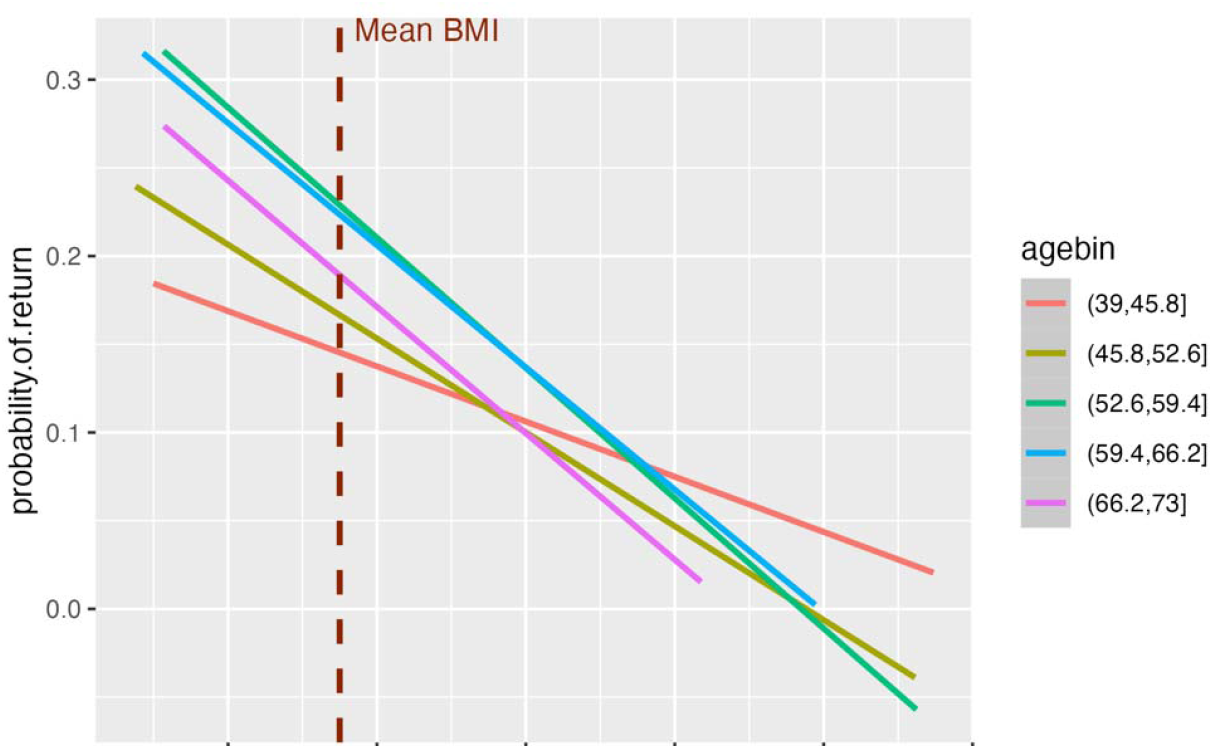
Evaluating the relationship between the level of BMI and probability of turning up to repeat clinic visit across age groups. The plot depicts the interaction between level of BMI and age on probability of returning to a repeat clinic visit. The x-axis depicts the level of BMI, and the y-axis depicts the probability of participants turning up to a repeat clinic assessment.

## Discussion

In this study, we conducted an exploratory analysis to investigate whether genetic associations with BMI and blood pressure traits (PP, SBP and DBP) exhibit age-dependent variation within the 40-69-year age range in the UK Biobank cohort. Specifically, we focused our attention on the feasibility of detecting age-varying SNP associations using an age-stratified GWAS framework. Given that access to individual-level data is often restricted in cohort studies, we have demonstrated an analytical framework which may have wide applicability for the comprehensive identification of age-varying genetic effects across the lifecourse using summary-level data (**Figure 1**).

The results of our analysis highlight distinct age-varying genetic profiles across the traits evaluated. For instance, a large proportion (47.7%) of our discovery set of genetic associations for PP had evidence for an age-varying effect as identified by our Inclusion 1 criterion. Notably, a similar profile was not observed for SBP (17.7%) or DBP (13.9%). Age-related changes in physiological blood pressure in individuals over 50-60 years old are characterized by a progressive increase in SBP, a stabilization or decline in DBP and consequent rise in PP (37). Age-associated arterial stiffness is a major contributor to the divergent trajectories of SBP and DBP in older people (38). By capturing the difference between SBP and DBP, PP is often used as a surrogate measure of arterial compliance and is an important predictor of adverse cardiovascular outcomes (39, 40). The enrichment of age interaction effects among discovery SNPs for PP relative to SBP and DBP aligns with the underlying relationship between PP and age. Furthermore, this supports the validity of the statistical approach applied here to identify context specific age-varying genetic effects. The trait dependent relationships with age produced in the present analysis were further supported by our sensitivity test where we modelled individual level SNP^*^Age interactions across the full age range, and similar evidence from the recent literature where a peak in the detection of SNP^*^Age interactions for PP relative to other traits have been described further (41).

This study provides a demonstration of the utility of the meta-regression framework to improve our understanding of the genetic architecture of complex traits. When comparing the relationship between GWAS and meta-regression estimates, in general the SNPs captured by our Inclusion 1 criterion (i.e. the set of discovery SNPs with evidence for an age-varying effect, P<0.05) had an overall attenuation in effect on BMI as age increased (**Figure 2**). The additional age-interacting SNPs identified by our Inclusion 2 criterion (i.e. the additional SNPs across the genome with evidence for an age-varying effect, P<5×10^-05^ ) represent a distinct cluster which similarly attenuated in effect on BMI with increasing age. In contrast, the opposite is observed for SNPs associated with PP identified using the Inclusion 1 and 2 criteria, which had an overall increasing effect trajectory on PP with increasing age. These patterns highlight differential influences of age on genetic associations detected within the 40-69-year age range evaluated for BMI and PP respectively and aligning with trends described previously in the recent literature (41).

Our results build on the existing literature where the detection of age-varying genetic associations across adulthood have been described, including those which have included the UKB cohort. For example, cross-sectional SNP^*^Age interaction analyses on a sample of 270,276 individuals in the UKB were conducted by Ao et al (42). They identified evidence of a small number of loci which attenuated in effect with increasing age for apolipoprotein B and triglycerides (including the APOE4 allele), although they did not detect strong interaction effects for other traits. More recently, Winkler et al describe SNP^*^Age interactions identified in a sample of 370,000 UKB participants for 8 traits (41). They report evidence for genetic effects on BMI and low-density lipoprotein cholesterol which attenuate with age, and substantially more for PP which tended to increase in effect with age. Importantly, top loci reported by Winkler et al with evidence for age-varying SNP effects on BMI (e.g. mapping to APOE) and PP (e.g. mapping to PIK3CG) were consistent with our results. Overall, our findings align with the literature indicating that the majority of SNP effects remain constant across the mid-late adulthood period of the lifecourse (1, 42). Though we have identified a subset of variants which had evidence for an interaction with age, the trend and the mechanism of the interaction is trait dependent.

While the previous literature has focused on the implementation of cross-sectional individual level SNP^*^Age interaction model, we have generated a resource where we have conducted complementary analyses, but which may also be used for analyses considering alternative models and assumptions. Furthermore, the summary level framework is an attractive model for initiatives aiming to consider age-varying effects across a wider range of the lifecourse by incorporating additional cohorts where summary statistics may be more readily shared. The age-stratified framework used was largely informed by Pagoni et al, where the detection of age-varying genetic effects by meta-regression was shown to perform best when the number of studies is high and the between study age-diversity is low (32). The major caveat of this approach is the impact of stratifying the sample population on the power to detect strong trait effects within each age-specific GWAS result. For this reason, we have applied relatively lenient significance thresholds within our inclusion criteria to explore candidate loci with suggestive evidence of age-dependent trait effects. When considering the detection of SNP^*^Age interaction effects within a cohort, the cross-sectional individual-level model remains a more highly powered approach.

We further explored the potential influence of SNP^*^Age interactions across the 40-69-year age range on exposure-outcome relationships estimated using MR. Our initial analysis of the relationships between each set of SNPs (discovery, Inclusion 1 and Inclusion 2) on cardiovascular and cardiometabolic outcomes by standard two-sample MR was primarily to provide a reference for the expected trait effects on each of the outcomes of interest. That said, MR estimates indicating strong evidence of an effect when instrumenting traits using our Inclusion 2 set of variants, for example, BMI on T2D and AF, and PP on T2D and PAD highlights that potential additional power may be gained by exploring the data for SNP^*^Age interactions genome-wide to yield relevant disease mechanisms which would otherwise be missed by the standard discovery GWAS.

In general, BMI instrumented using the discovery GWAS SNPs strongly predicted elevated risk of developing each of the cardiovascular and cardiometabolic outcomes in our standard MR analysis, while the SNPs with evidence for an age-varying effect on BMI identified by our Inclusion 1 and 2 criteria were most relevant for AF and T2D risk. The results of our age-stratified standard MR analyses emphasizes the stronger effect of higher BMI at older age (68-69 years) on lifetime risk of disease compared to BMI measured earlier in life (40-41 years). While the results of our modified ivw analysis indicated that the SNP^*^Age interaction has a diminishing effect on the rate of change in BMI due to genetic effects on the relationship between BMI and the cardiovascular and cardiometabolic outcomes analyzed. This pattern was observed for BMI when estimated using both the discovery and Inclusion 1 set of SNPs on PAD, HF, CHD, AF and T2D, and may suggest that the interaction with age is most pronounced on variants with the largest trait effect. This aligns with previously described examples where BMI-associated SNPs have been shown to elicit a stronger trait effect earlier in the lifecourse (43-46). Taken together, these results highlight dynamic changes between the genetic and environmental influences on population level BMI across the lifecourse. This suggests a mechanism where individuals with higher genetic predisposition to BMI may experience longer exposure to BMI throughout the lifecourse, while individuals with lower genetic predisposition experience more environmentally driven change in BMI in older adulthood. This is also demonstrated by longitudinal studies, where predictive models of weight gain were only improved by polygenic risk score inclusion in early adulthood but not in middle and later aged adults (47). Other recent work has shown that for some quantitative traits, heritability decreases with age relative to the accumulative exposure to environmental variance (48). This is particularly relevant for BMI, where environmental factors have been implicated as the main driver of longitudinal trait change relative to genetic factors across older ages of adulthood (49, 50). Additionally, the emergence of the obesogenic environment over the late 20^th^ century has been shown to drive stronger gene^*^environment interaction effects on BMI influencing generational differences in the level of BMI (51, 52). Given the cross-sectional design of UKB we could not estimate generational effects in the present study, though this may be an important consideration for future work incorporating additional cohorts.

In contrast, our analyses also highlighted an example where older age in the population may be important for the detection of genetic predictors for the exposure. Results of the MR analysis of PP instrumented using our discovery, Inclusion 1 and Inclusion 2 SNP sets, showed evidence of an effect on outcomes such as stroke, HF and PAD. The results of the modified ivw analysis support a relationship where the variation in genetically predicted PP with age maintains a strong effect on the risk of developing stroke, PAD, HF and CHD. Importantly, similar results were observed when instrumenting PP with the Inclusion 2 SNPs in the modified ivw framework for stroke, PAD and HF lending further strength to the proposed analytical framework to yield further insight on genetic factors underlying disease progression. Cross-comparison with the results of the same analysis but for SBP emphasizes that the detection of putatively functional age-interaction effects is dependent on relevance to the trait. There was little evidence to suggest effects of age on the relationship between genetically predicted SBP and the secondary cardiovascular outcomes assessed in our modified ivw analysis, except between our Inclusion 1 set and PAD only. This is likely attributable to the exaggerated widening in PP relative to the increase in SBP across the same age range. Further investigation of SNP^*^Age interaction associations for PP may help identify biological mechanisms implicated in the progression of these secondary outcomes.

We have conducted detailed investigations on the potential influence of selection on our ability to detect reliable age-varying genetic associations within the age-stratified framework. Importantly, genetic determinants of lipid and cardiovascular outcomes have been demonstrated to have a detrimental effect on survival (28). As such it is possible that a higher prevalence of comorbidities such as CHD and stroke with increasing age could have an effect on the detection of SNP^*^Age interactions. Our simulation study showed that age-varying genetic effects could be induced in the presence of interaction effects between the trait of interest and age on the likelihood of participating in the study. However, such effects would be expected to apply across all SNPs associated with the trait (i.e. all discovery SNPs). For BMI, SBP and DBP only a limited proportion (∼10-15%) of the discovery SNP set had evidence of a SNP^*^Age interaction at a nominal p-value threshold (P<0.05), suggesting that the SNP^*^Age interactions identified in our study are unlikely to have been induced by selection. For PP a higher proportion of the discovery SNPs showed age-varying genetic effects. While SNPs with evidence for an age-varying effect on BMI predominantly exhibited an attenuation in effect, we identified evidence for an increasing interaction effect of rs429358 (T allele) on BMI with age according to our Inclusion 2 criteria. The C allele of rs429358 is an established predictor of Alzheimer’s disease risk (22, 53) and early mortality (54). An examination of SNP^*^Age interaction effects by Winkler et al describe in a sensitivity analysis that the rs429358 SNP presented evidence for a putative age-dependent effect on selection in UKB. rs429358 was the only variant identified in their analysis which presented evidence for a residual association with age, further indicating that selection effects were not pervasive among SNPs with evidence for an age interaction. Previous efforts to identify SNPs with strong survival effects have been investigated in cohorts beyond age 70 and identified only a small number of variants (notably at the APOE and CHRNA3 loci) (28). Lastly, the evaluation of the relationship between age and BMI on the probability of returning to repeat assessments in the UKB (**Figure 7**) illustrates that the negative association between BMI and the probability of returning is largely consistent across age groups, except that it was perhaps weaker in the youngest age group. Taken together these analyses support that the age-varying genetic effects identified overall are unlikely to be biased by age varying selection effects.

This work has a number of limitations. While the age-stratified summary data generated in this study may facilitate the identification of age-varying associations without imposing a linear constraint on the data, the meta-regression framework implemented in the present study only considered linear relationships with age. It is possible that more acute age effects may underlie some traits in the population. Investigating these would require more complex modelling, outside the scope of the present study. We note that UKB participation was voluntary and participants have on average healthier lifestyles, higher levels of education and better health than the general population (55-57). This healthy-participation bias has been further shown to distort the identification of some genetic associations (58). Given this selection, the detection of SNP^*^Age interaction effects on general health traits in UKB by age 69 may be diminished relative to the general population. Additionally, our analyses were limited to the inclusion of participants of European descent, as such these results are not necessarily generalizable across other ancestries.

In conclusion, we have comprehensively evaluated genetic associations across the 40–69 age range in the UKB cohort at 2-year intervals for BMI and blood pressure traits (PP, SBP, DBP). All GWAS summary data derived in this study will be made readily accessible (upon acceptance of the manuscript for publication) to allow further exploration of SNP^*^Age interactions identified using the age-stratified framework. To uncover genetic variants modified by age, SNP estimates were systematically incorporated into a meta-regression framework, shedding light on trait-dependent patterns of age interaction effects. Furthermore, we demonstrate within an MR and modified ivw framework how the identification of age-varying effects may provide extended context to inform the interpretation of genetic predictors identified by conventional cross-sectional GWAS analyses for improved causal inference.

## Methods

### Study Population

The UKB is a highly phenotyped prospective cohort study encompassing >450,000 participants aged 40-69 years at recruitment (12, 54). GWAS associations were initially generated for each trait on imputed data for all individuals who passed our selection filters (see below ‘GWAS’ section for detail on the filters applied) aged 40-69 years (N=463,005), which will herein be referred to as the ‘discovery’ dataset. A small number of participants who fell outside this age range were excluded due to low representative power (number of individuals excluded: 2,114). Participants were next stratified into one of 15 mutually-exclusive two-year age strata and GWAS were conducted within each stratum for each trait. As participant age in the UKB study is skewed towards the upper range, to account for the differential sample size between strata, we applied a maximum sample size cut-off (matching the sample size in stratum 7 (52-53 year olds), N=26,059) and this many individuals were randomly sampled from strata with a larger sample size.

GWAS analyses were conducted using individual level data obtained from the UKB cohort study for 4 complex traits (BMI, PP, SBP, DBP,). PP was calculated by subtracting DBP from SBP. Blood pressure traits were obtained from the mean of two measures, and were adjusted for anti-hypertensive medication by adding 15mmHg to SBP and 10mmHg to DBP for individuals who self-reported use of any anti-hypertensive medication (17, 59). The following UKB datafields were obtained for blood pressure measures; SBP: 4080-0-0, 4080-0-1; DBP: 4079-0-0, 4079-0-1. To identify participants taking anti-hypertensive medication the UKB datafields 6177-0.0 (male participants) and 6153-0.0 (female participants) were extracted. Across our sample population we identified 87,838 participants taking anti-hypertensive medication (20.04%). It should be acknowledged that this transformation has several limitations. Firstly, the data is unadjusted for other medication use, which may also influence blood pressure. Secondly, the transformation is based on self-reported medication use (which may be less reliable than other medical records) and applied as a constant which may not provide an accurate representation of the impact of anti-hypertensive drugs. BMI measurements were obtained from UKB datafield 21001-0-0, where BMI values were constructed from height and weight measured during the initial assessment center visit. Mean phenotypic values and sample sizes in each age stratum are provided in **Table S22**.

### GWAS

The full UKB data release contains the cohort of successfully genotyped samples (n=488,377). 49,979 individuals were genotyped using the UK BiLEVE array and 438,398 using the UK Biobank axiom array. Pre-imputation QC, phasing and imputation are described elsewhere (15). Genotype imputation to a reference set combining the UK10K haplotype and HRC reference panels (60) was performed using IMPUTE2 algorithms (61). Quality control (QC) and filtering of the UKB genetic dataset was conducted by R. Mitchell, G. Hemani, T. Dudding, L. Corbin, S. Harrison, L. Paternoster as described in the published protocol (doi:10.5523/bris.1ovaau5sxunp2cv8rcy88688v). The genetic data provided by UKB has been filtered to include 12,370,749 SNPs. The sample was restricted to individuals of ‘European’ ancestry as defined by an in-house k-means cluster analysis performed using the first 4 principal components provided by UK Biobank in the statistical software environment R. A full description of the analytical pipeline and QC has been described previously (62, 63). The present analyses are limited to the inclusion of participants of European ancestry, which is the major ancestry group in the UKB. While other ancestry groups are present in the UKB, because we are stratifying the study population by age all other ancestry groups had relatively low overall sample sizes which led to very small available sample sizes for each age-strata.

GWAS were conducted using a linear mixed model (LMM) association method as implemented in BOLT-LMM (v2.3) (64) with adjustment for age, sex and genotype-chip, using the default analysis parameters defined in an in-house MRC-IEU UKB GWAS pipeline (doi: https://doi.org/10.5523/bris.pnoat8cxo0u52p6ynfaekeigi). In brief, to model population structure 143,006 directly genotyped SNPs were used, obtained after filtering on MAF > 0.01; genotyping rate > 0.015; Hardy-Weinberg equilibrium p-value < 0.0001 and LD pruning to an r^2^ threshold of 0.1 using PLINKv2.00. BOLT-LMM association statistics are on the linear scale. As such, test statistics (betas and their corresponding standard errors) were transformed to log odds ratios and their corresponding 95% confidence intervals on the liability scale using a Taylor transformation expansion series (65). The proportion of trait variance explained by genetic variants in each GWAS analysis was estimated by the infinitesimal model used by the BOLT-LMM software. A description of the age strata and their respective sample sizes is provided in **Table S18**. All summary level results generated in these analyses will be made available online upon acceptance of this manuscript for publication. Independent genome-wide SNP associations were identified via linkage-disequilibrium (LD)-clumping using PLINK (v1.9) (66). A reference panel comprising data on 10,000 unrelated individuals of European ancestry in the UKB (67) was incorporated to identify independent SNPs based on r^2^ <0.01 and a genome-wide significance threshold (P<5×10^-08^ ). Genetic variants identified at these thresholds in our non-age stratified cross-sectional GWAS analyses are referred to as the “discovery” set of variants.

### SNP^*^Age interaction analysis in age-stratified summary data by meta-regression

To formally identify SNPs with evidence of an age interaction on each of the specified traits, we used meta-regression using the ‘metafor’ package in R. For each SNP, 15 datapoints capturing the respective age-specific GWAS estimates derived in each stratum were fitted to a linear fixed effects meta-regression, with mean age in each bin as the exposure. As such, our selection criteria for SNP^*^Age interactions were based on: (i) the subset of ‘discovery’ SNPs with evidence of an age-varying effect in the meta-regression (P<0.05), and (ii) additional independent SNPs identified genome-wide with evidence of an age-varying effect in the meta-regression (P<5×10^-05^, r^2^ <0.01). Herein, the SNPs identified according to these criteria are referred to as the ‘Inclusion 1’ and ‘Inclusion 2’ SNP sets, respectively. In our Inclusion 1 set, we used the power of the full cohort and conventional genome-wide significance thresholding (P<5.10^-08^ ) to consider the set of independent (r^2^ <0.01) trait-associated variants which also showed evidence for an age-varying effect (at P<0.05). In our Inclusion 2 criteria, we applied strong thresholds for significance and independence to uncover any additional loci with suggestive evidence for an age-varying effect. In subsequent analyses, we use the SNPs identified by our inclusion criteria to assess, broadly, the impact of age at exposure measurement on genetically proxied exposure-outcome relationships. We highlight top effects which survived more stringent Bonferroni thresholding within each group, but maintained the more relaxed threshold within our inclusion criteria. Independent effects were detected by LD-clumping using PLINK 1.9 (66). Plots for visual comparison were generated using the ‘ggplot2’ package in R.

### SNP^*^Age interaction analysis in individual level data

For comparison, SNP^*^Age interactions were evaluated in a non-age stratified setting using individual-level data for all individuals aged 40-69 years for each trait (blood pressure traits, N=433,481; BMI, N=458,360). genome-wide SNP^*^Age interaction analyses were performed on the same quality controlled and filtered UKB genotype dataset as described above. The genetic (G) main effect and G^*^Age (interaction effect) were modelled using ‘fastGWA-GE’ mixed linear model from ‘GCTA’ with adjustment for sex, genotype chip and the first 10 genetic principal components. In the analysis, rare variants (MAF <0.01%) were excluded and only variants which had a high imputation quality (INFO >0.8) were retained. Genome-wide significant main and interaction effects were detected by LD-clumping using PLINK (1.9) (66), using the UKB reference panel of 10,000 individuals as described previously (67). We first report the number of loci with a main genetic (trait) effect in the analyses (P<5×10^-08^, r^2^ <0.01). We then highlight the proportion of these SNPs which had evidence of an interaction (G^*^E) at P<0.05 (analogous to the inclusion 1 criterion in our primary analysis). We next focus on the 1 degree of freedom test of the SNP^*^Age interaction. We highlight the number of loci which had evidence for an interaction effect at P<5×10^-05^ and r^2^ <0.01 (i.e. analogous to our Inclusion 2 criterion), and further highlight which SNPs survived standard genome-wide significance thresholding (genome-wide significance (P<5×10^-08^ ).

### Mendelian randomization analysis of age-varying genetic effects

To investigate the influence of age on exposure-outcome relationships estimated using genetic methods, we conducted a series of MR analyses on 6 cardiovascular related outcomes: T2D, stroke, PAD, HF, CHD and AF. An overview of all outcome datasets is provided in **Table S23**. The exposure-outcome relationships were first estimated by ivw summary data MR analysis using the ‘TwoSampleMR’ package in R separately for each set of SNPs (discovery, Inclusion 1 and Inclusion 2). The aim of this analysis is to provide a reference for how exposures predicted using the overall (discovery) and age dependent (Inclusion 1 and 2) instruments perform in the model. The results of MR analyses using the discovery set of variants illustrate the effect of the genetically derived exposure on the outcome per standard deviation increase in the exposure. The results of analyses using the Inclusion 1 subset of SNPs demonstrate whether the overall relationship is replicated by SNPs with an age-dependent effect. The results of analyses using Inclusion 2 SNPs suggest putative age-dependent exposure-outcome relationships which were not captured using the standard discovery SNPs alone. The incorporation of these sets of SNPs into the MR model is helpful in illustrating the potential impact of age on genetically derived exposure-outcome relationships, though we do not interpret these as precise causal estimates. We applied MR robust methods (weighted median and MR Egger) as a sensitivity check.

To provide an indication of how the exposure-outcome relationships vary depending on the age at which the exposure is measured, further age-stratified MR analyses were conducted. In these analyses, exposure-outcome relationships were estimated using effect estimates derived in the youngest (40-41 year) and oldest (68-69 year) strata respectively using the discovery set of SNPs.

### Estimating the effect of age on the genetic liability underlying the exposure-outcome relationships

To assess the effect of the SNP^*^Age interaction on genetically predicted exposure-outcome relationships, we modified the standard ivw MR analysis to explicitly consider the SNP^*^Age interactions. To achieve this, we incorporated the meta-regression effect estimates as proxy “exposures” in a summary data MR analysis (as opposed to the GWAS SNP-exposure association estimates). Results from this analysis were the effect estimates (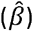) obtained from ivw estimation of the summary data model;

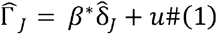

Where the estimated values of the SNP-outcome association 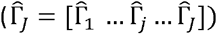 were obtained from;

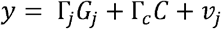

For each SNP *j, C* is a set of covariates including sex, genotype chip, age and the first 10 principal components of ancestry, Γ_*C*_ is the effect of those covariates on *y*. The estimated values of the change in the SNP-exposure association over increasing age 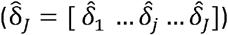 was obtained from a two step estimation. Firstly, the estimated association between each SNP *j* and the exposure is obtained for each age group *a* from;

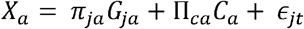

Where *X*_*a*_ and *X*_*ja*_ are the subsets of *X* and *G* for individuals in age group *a* and *C*_*a*_is the same set of covariates as used previously, Π_*ca*_ is the effect of those covariates on *X* for age group *a*. Secondly the effect of age on the estimated association between the SNP and the exposure (i.e. 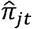 for each SNP (*j*) was then obtained from the regression;

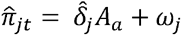

Where *A*_*a*_ is the mean age in age group a. This estimates the effect of the rate at which the SNP-exposure association changes with increasing age on the risk of the outcome. We refer to the estimation of 1 as a Modified ivw analysis, however we do not interpret the estimates obtained as causal effect estimates but instead use this framework to identify the relationship between the age-varying effect of the SNPs and disease outcomes. Modified ivw analyses were conducted using the SNPs identified in the discovery, Inclusion 1 and Inclusion 2 sets as above.

### Simulation study to evaluate potential bias due to selection in the age-stratified framework

We conducted a simulation study to estimate the potential for selection bias in study participation to induce an age-varying genetic effect when the true effect of the SNP on the trait is constant over different ages (36). We generated a dataset including a trait of interest *X*, age *A* generated as a uniformly distributed variable, a single genetic variant *G*, generated from a binomial distribution with minor allele frequency of 0.4, and a normally distributed unobserved confounder *U*. The relationship between these variables was given by;

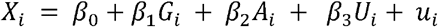

Where *u* is a normally distributed random error term. 50% of the generated individuals were considered ‘observed’ and the rest of the dataset was set to missing for all observations. Selection was determined by the expression;

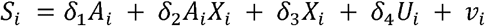

Where *u*_*i*_ is a normally distributed random error term. The 50% of individuals with the highest values of S were set to missing. The observed dataset was then stratified into 4 age categories and the association between the genetic variant and the trait estimated in each group separately using the regression model;

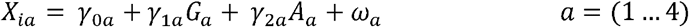

We varied the values of *δ*_1_, *δ*_2_ and *δ*_3_ to assess the impact of different selection mechanisms in the bias in the estimated value of *γ*_1*a*_ across the different age categories. This was repeated twice, once for a model with an effect of on (i.e. *β*_1_ ≠ 0 and once for a model with no effect of on (i.e. *β*_1_ = 0 . Bias in the estimated effects across the different parameters considered are given in **Figure 5**. The proportion of the variation in missingness explained by each of and A across the levels of each value of *δ* _1_, *δ*_2_ and *δ*_3_ is given in **Table S24-S25**.

### Empirical investigation of sources of selection with increasing age

To investigate trait and age effects on selection, we used the subset of the UKB who received an invitation to attend a repeat clinic visit (datafield: 110008). Out of 101,022 individuals, 20,177 attended the repeat clinic (19.97%). Using BMI as exemplar, we evaluated the average association between BMI and the probability of attending the repeat clinic across 5 age groups. This was achieved by fitting a linear model that included BMI as a predictor, along with dummy variables for each age category and interaction terms between BMI and these age categories. The fitted values were plotted, indicating the predicted probability of attending the clinic according to age group and BMI. Further analyses examined using logistic regression the relationship between the trait^*^age interaction term and the probability of attending the repeat assessment clinic across the overall age-range in the sample, and within 5 age groups for each trait (BMI, PP, SBP, DBP).

## Data Availability

All data generated as part of this study are currently under restricted access on Zenodo. Access to this dataset will be made publicly available upon acceptance of this work for publication.

## Acknowledgements

This research has been conducted using the UK Biobank Resource under Application Number 81499. This work was funded by the UK Medical Research Council (MRC) as part of the MRC Integrative Epidemiology Unit (MC_UU_00032/1 to GH and GDS, and MC_UU_00032/2 to KT). ES is supported by the MRC (UKRI077). The funders had no role in study design, data collection and analysis, decision to publish, or preparation of the manuscript.

## Author Contributions

GML: Formal Analysis, Investigation, Visualization, Writing – Original Draft Preparation, PP: Resources, Writing – Review & Editing, GMP: Writing – Review & Editing, DC: Writing – Review & Editing, TGR: Resources, Writing – Review & Editing, KT: Writing – Review & Editing, GH: Resources, Writing – Review & Editing, GDS: Conceptualization, Writing – Review & Editing, ES: Conceptualization, Supervision, Methodology, Formal Analysis, Writing – Review & Editing

## Supporting Information (list of legends)

### Supplementary tables

Table S1: Summary of lead genetic variants identified in individual age-strata for BMI

Table S2 Summary of lead genetic variants identified in individual age-strata for PP

Table S3: Summary of lead genetic variants identified in individual age-strata for SBP

Table S4: Summary of lead genetic variants identified in individual age-strata for DBP

Table S5: Inclusion 1 set: Subset of BMI discovery SNPs with evidence for an age varying genetic effects (Meta-regression P<0.05)

Table S6: Inclusion 1 set: Subset of PP discovery SNPs with evidence for an age varying genetic effects (Meta-regression P<0.05)

Table S7: Inclusion 1 set: Subset of SBP discovery SNPs with evidence for an age varying genetic effects (Meta-regression P<0.05)

Table S8: Inclusion 1 set: Subset of DBP discovery SNPs with evidence for an age varying genetic effects (Meta-regression P<0.05)

Table S9: Inclusion 2 set: Additional SNPs identified with evidence for an age varying genetic effect on BMI (Genome-wide meta-regression P<5×10-05, r2<0.01)

Table S10: Inclusion 2 set: Additional SNPs identified with evidence for an age varying genetic effect on PP (Genome-wide meta-regression P<5×10-05, r2<0.01)

Table S11: Inclusion 2 set: Additional SNPs identified with evidence for an age varying genetic effect on SBP (Genome-wide meta-regression P<5×10-05, r2<0.01)

Table S12: Inclusion 2 set: Additional SNPs identified with evidence for an age varying genetic effect on DBP (Genome-wide meta-regression P<5×10-05, r2<0.01)

Table S13: SNPs with evidence of a SNP^*^Age interaction effect on BMI from our individual level analysis.

Table S14: SNPs with evidence of a SNP^*^Age interaction effect on PP from our individual level analysis.

Table S15: SNPs with evidence of a SNP^*^Age interaction effect on SBP from our individual level analysis.

Table S16: SNPs with evidence of a SNP^*^Age interaction effect on DBP from our individual level analysis.

Table S17: Standard ivw MR analysis of exposure variables instrumented separately using discovery, inclusion 1 and inclusion 2 SNPs

Table S18: Sensitivity MR analysis of exposure variables instrumented separately using discovery, inclusion 1 and inclusion 2 SNPs using MR robust methods

Table S19: Results of age-specific ivw MR analyses of exposure variables instrumented using age-stratified GWAS estimates

Table S20: Modified ivw analyses of exposure variables instrumented separately using discovery, inclusion 1 and inclusion 2 SNPs

Table S21 Evaluation of the relatoinship between trait^*^age interaction effects on the probability of UKB participants to return to repeat clinic assessments

Table S22 A description of the age strata and their respective sample sizes used in all GWAS analyses.

Table S23 Detail of outcome datasets used in Mendelian randomization analyses

Table S24 Selection simulation 1: Outline of the proportion of the variation in missingness explained by each of G and A across the levels of each value of *δ*_1, *δ*_2 and *δ*_3

Table S25 Selection simulation 2: Outline of the proportion of the variation in missingness explained by each of G and A across the levels of each value of *δ*_1, *δ*_2 and *δ*_3

### Supplementary material

Supplementary Note 1: Descriptive overview of effect size differences in our age-stratified GWAS results.

Figure S1 Comparison of GWAS effect estimates between Stratum 1 (40-41 years) and Stratum 8 (54-55 years) for BMI.

Figure S2 Comparison of GWAS effect estimates between Stratum 1 (40-41 years) and Stratum 15 (54-55 years) for PP.

Figure S3 Comparison of GWAS effect estimates between Stratum 1 (40-41 years) and Stratum 8 (54-55 years) for SBP.

Figure S4 Comparison of GWAS effect estimates between Stratum 1 (40-41 years) and Stratum 8 (54-55 years) for DBP.

Figure S5 Comparison of GWAS effect estimates between Stratum 1 (40-41 years) and Stratum 15 (68-69 years) for BMI.

Figure S6 Comparison of GWAS effect estimates between Stratum 1 (40-41 years) and Stratum 15 (68-69 years) for PP.

Figure S7 Comparison of GWAS effect estimates between Stratum 1 (40-41 years) and Stratum 15 (68-69 years) for SBP.

Figure S8 Comparison of GWAS effect estimates between Stratum 1 (40-41 years) and Stratum 15 (68-69 years) for DBP.

Figure S9 Comparison of GWAS effect estimates between Stratum 8 (54-55 years) and Stratum 15 (68-69 years) for BMI.

Figure S10 Comparison of GWAS effect estimates between Stratum 8 (54-55 years) and Stratum 15 (68-69 years) for PP.

Figure S11 Comparison of GWAS effect estimates between Stratum 8 (54-55 years) and Stratum 15 (68-69 years) for SBP.

Figure S12 Comparison of GWAS effect estimates between Stratum 8 (54-55 years) and Stratum 15 (68-69 years) for DBP.

Figure S13 Scatter plot depicting the relationship between SNP-trait effect estimates derived by GWAS and SNP^*^age interaction effects derived by meta-regression analysis for (i) SBP and (ii) DBP.

Supplementary Note 2: Summary of overlapping loci in sensitivity analysis

Table 1 Summary of the number of exact SNPs identified in the sensitivity analyses using individual level data with the primary analysis.

Supplementary Note 3: The results of age-stratified MR analyses

Figure S14 Results of age-specific standard MR analyses for BMI, PP, SBP and DBP.

Figure S15 Standard MR estimates of (i) the effect of genetically proxied SBP measured at 40-69 years old on six cardiovascular outcomes and (ii) modified ivw estimates depicting the change in the effect of genetically proxied SBP on those outcomes per additional year of age at measurement.

Figure S16 Standard MR estimates of (i) the effect of genetically proxied DBP measured at 40-69 years old on six cardiovascular outcomes and (ii) modified ivw estimates depicting the change in the effect of genetically proxied DBP on those outcomes per additional year of age at measurement.

